# Pain on the Street: Implementation of a Field-Based Pain Response for People Experiencing Homelessness

**DOI:** 10.64898/2026.08.13.26360409

**Authors:** Sarah Jean Valliant, Manav Joseph, Misha Sethi Sharma, Gainu Phela Durosinmi, Dayla Fitts, Sophia Tran, Abigail Noguez Gonzalez, Sonya Kothari, Trevor Anderson, Riya Ralh, Angelina Ka Lee, Samantha S. Parton, Fazila Shirinzada, Carole Kulik

## Abstract

2.

**Background:** Homeless individuals are disproportionately affected by limited access to primary care and pain management services. While community-based organizations frequently conduct outreach, few have ethical, standardized, or replicable methods for assessing pain and distributing referrals in field settings. Existing models range from passive, meals-only outreach, to costly mobile clinics with limited reach. This leaves a critical gap that a low-resource, agile framework is designed to address.

**Objective:** The aim of this quality-improvement initiative was to implement and iteratively refine a standardized, field-based pain assessment and response pathway for adults experiencing homelessness and to evaluate its feasibility, fidelity, safety, and operational barriers during routine outreach.

**Methods:** A cross-sectional quality improvement needs assessment was conducted during homeless outreach activities in San Francisco and Sacramento using convenience sampling. The intervention framework incorporated volunteer training, cognitive capacity screening, verbal informed consent, vital sign collection, and predefined criteria for emergency escalation. Participants were unsheltered adults with adequate decisional capacity to provide voluntary informed consent.

**Results:** The dataset included 193 encounters, with valid pain scores for 175 participants. Mean pain was 3.79 ± 2.78, with a median of 3. Cold packs were used for acute discomfort and foot or ankle pain, while hot packs were used for joint pain. Some participants declined comfort measures. No supply shortages, referral confusion, or emergency-escalation delays were documented. Telephone access remained a barrier to referral completion.

**Conclusion:** This framework demonstrates a replicable and ethically grounded model for field-based pain assessment and referral during homeless outreach. The pathway was feasible and safe to implement, expanded care options beyond default emergency-department referral by directing stable nonemergent pain toward primary care, and identified limited telephone access as a major barrier to completing follow-up.

## Introduction

### 3. Problem Description

Individuals experiencing unsheltered homelessness encounter higher rates of acute and chronic pain while facing difficulty obtaining outpatient care may disproportionately rely on emergency medical resources for episodic pain care when primary care is inaccessible or difficult to maintain.

Despite the presence of advanced healthcare infrastructure and specialized outreach efforts, providing adequate medical care for unhoused individuals continues to be a challenge. Structural barriers, including limited access to communication, lack of a permanent address, lack of access to sanitation, private facilities, and limited mobility, inhibit preventive services and continuous care from reaching unhoused populations [10,11].

Many of these barriers, including challenges in meeting subsistence needs and primary care, have been documented [11, 14]. Due to these obstacles, essential services such as diagnosis and treatment are delayed or unavailable, oftentimes resulting in unhoused individuals only receiving medical attention when experiencing medical emergencies.

In efforts to address overdependence on emergency medical services, programs and organizations, including the San Francisco Fire Department’s Homeless Outreach and Medical Emergency (HOME) Team, have been established to support unhoused individuals who periodically need medical treatment and assistance [15]. Many medical outreach models and organizations focus on addressing emergency or substance use crises, leaving less support and guidance for routine, nonemergent care.

The scope of the problem extends beyond an increased burden on emergency medical resources and is also characterized by challenges in determining appropriate levels of care, lack of accessibility to primary care options and immediate nonpharmacologic support, and barriers to obtaining referrals due to factors such as limited access to a phone and unreliable transportation.

Despite the presence of specialized outreach, ensuring medical care reaches unhoused individuals remains a complex challenge. Even when resources might be available, mistrust between unhoused individuals and healthcare institutions due to previous experiences of discrimination and mistreatment often keep unhoused individuals from pursuing medical services.

In an effort to gain insight on the issue of acute pain faced by individuals experiencing unsheltered homelessness, the Valliant Foundation led outreach missions in Sacramento and San Francisco to record pain scores and identify emergencies. During these missions, volunteers did not have a standardized method for responding to stable nonemergent pain. However, based on repeated reports of acute and chronic pain, it became evident that standardized pain-response pathways are needed.

### 4. Available Knowledge

Existing literature documents various factors responsible for health disparities experienced by unhoused individuals, such as institutional mistrust, food insecurity [4], and environmental stress [3]. As a result of disparities in access to healthcare in San Francisco, unhoused individuals are disproportionately subject to experiencing severe pain. Beyond this, no consistent, field-adapted framework focused on preventing and treating severe pain is thought to exist.

Implementation frameworks focused on health equity identify and highlight the impact of structural barriers experienced by unhoused individuals on ensuring healthcare accessibility and delivery. These frameworks emphasize the importance of developing screening approaches that focus on addressing historical systems of disadvantage, environmental factors, and participant vulnerability.

Specialized outreach programs focused on providing health education and basic resources are essential to addressing disparities in healthcare for unhoused individuals [2], [7]. Current data primarily reflects unhoused populations with access to shelters or outpatient clinics, omitting unhoused individuals who are the most marginalized and for whom traditional healthcare strategies are financially and logistically impractical. Beyond this, many specialized outreach programs operate without standardized and ethically reviewed protocols. As a result, adequate capacity assessment, informed consent, and proper emergency escalation are not guaranteed when providing medical services [17].

To address this concern, the Valliant Foundation designed a Quality Improvement (QI) framework focused on screening unhoused individuals in San Francisco for severe pain. A replicable methodology with details on emergency protocols, ethical supervision, and consent screening developed to improve service quality and outreach safety is presented in this paper.

### 5. Rationale

This initiative built upon an existing ethically reviewed outreach protocol. The current quality-improvement intervention focused specifically on standardizing the assessment and response to pain through a tiered referral pathway, optional nonpharmacologic comfort measures, and expanded resource navigation.

Although the existing protocol routinely identified participants experiencing acute and chronic pain, it did not provide volunteers with a standardized process for determining the appropriate level of care or selecting supportive comfort measures after pain was identified. Consequently, referral decisions and nonpharmacologic interventions depended largely on individual volunteer judgment.

The intervention standardized field-based on pain management by combining structured pain assessment with referral recommendations based on clinical urgency. Participants with emergency warning signs were offered immediate emergency assistance, severe nonemergent pain prompted same-day urgent care referral, and stable pain was directed to primary care of community clinics. Optional hot and cold packs and individualized resource navigation were incorporated to provide temporary comfort and reduce barriers to follow-up.

Consistent with quality-improvement methodology, this initiative emphasized implementation and iterative refinement of an operational outreach protocol rather than the development of research questions characteristic of Participatory Action Research or Community Based Participatory Research frameworks.

### 6. Specific Aims

This quality-improvement initiative introduced a tiered pain-referral pathway and standardized hot-and cold-pack interventions. The primary aims were to assess the feasibility and fidelity of directing participants to primary care, urgent care, or emergency services based on pain severity and warning signs; to provide appropriate low-barrier comfort measures; and to promote primary care for nonemergent pain while preserving immediate escalation for medical emergencies.

Consistent with established QI practice, community engagement informed referral pathways and field procedures rather than hypothesis generation or study governance. The manuscript follows the SQUIRE 2.0 framework to support transparency, reproducibility, and dissemination of operational lessons learned, rather than to present hypothesis-driven or theory-generating research [20].

## Methods

### 7. Context

The assessment was conducted from November 2024 to June 2026 during routine outreach activities in San Francisco and Sacramento. Outreach in San Francisco occurred in the Tenderloin, Inner Mission, and Embarcadero neighborhoods, while Sacramento outreach focused on the River District and surrounding areas. These service areas are characterized by high rates of housing instability, population transiency, limited access to healthcare, and significant socioeconomic challenges, making them important locations for community-based outreach and preventive health initiatives.

Typical outreach missions lasted approximately four hours and were staffed by teams of six to twelve trained volunteers. During the fall, winter, and spring, outreach missions were conducted approximately every other week, while summer missions were held monthly. The same standardized questionnaire, health screening protocol, and informed consent process were used at both the San Francisco and Sacramento sites to ensure consistency in data collection.

Participants at all outreach locations were offered the same core resources, including food and water, hygiene supplies, clothing, information on emergency shelters, housing assistance programs, mental health counseling, food assistance services, and guidance on obtaining government-assisted cell phones to improve access to healthcare and social services. Following implementation of the health screening protocol, hot and cold packs were also made available at outreach missions during June. Data collection was limited to daylight hours and favorable weather conditions to maximize volunteer safety, operational efficiency, and participant engagement.

### 8. Interventions

This Operational Needs Assessment was developed to guide nonprofit resource allocation and outreach operations rather than to serve as a formal human-subjects research study. Individuals requiring medical follow-up were referred through location-specific emergency departments, same-day urgent care clinics, federally qualified health centers and free clinics, behavioral health services, and public-benefit programs according to the urgency of their needs.

Participants reporting pain were assessed using standardized referral criteria based on pain severity and the presence of emergency warning signs (Table 4). Emergency findings superseded numerical pain scores when determining the appropriate level of care. When clinically appropriate, participants were offered optional hot or cold packs as temporary nonpharmacologic comfort measures and instructed on safe use.

### 9. Study of the interventions

Factors of ethical constraints, feasibility, and operational saturation determined sample size for the study, instead of statistical power calculations. For the study, the outreach program was conducted with strict capacity assessment, daylight exclusive engagement, and finite personnel. To prevent duplicate participation, volunteers verbally screened each individual by asking if they have taken a survey before during any prior outreach. Individuals who had previously participated were excluded from the study and data collection. Although they were not enrolled in the study, those individuals were still provided with the standard outreach resources including social support, resources, and health screening. The hot/cold-pack intervention was only provided during the month of June. Participants not encountered during the month of June received the standard outreach protocol interventions, but were not eligible to receive the hot/cold-pack intervention.

Implementation was evaluated across fidelity, feasibility, acceptability, and safety domains. Fidelity reflected adherence to required assessment and documentation procedures; feasibility captured operational barriers affecting protocol delivery; acceptability reflected participant acceptance or refusal of offered interventions; and safety included recognition and timely escalation of emergency findings or hazards.

Because this was a descriptive quality-improvement initiative, no inferential power calculation was performed, and the project was not designed to determine causal effects on pain, referral completion, healthcare utilization, or long-term outcomes.

Balancing measures were implemented to identify unintended burdens related to the implementation of the outreach protocol. Implementation was not noted as interfering with participant encounters. Cold packs were provided for acute or activity-related discomfort, including localized swelling, minor soft-tissue pain, and foot or lower-extremity pain associated with prolonged walking or standing. Hot packs were used primarily for musculoskeletal discomfort, including joint pain, muscle soreness, stiffness, and chronic or activity-related aches when heat was considered appropriate under the outreach protocol.

Clinical referrals were provided when necessary according to the outreach protocol. Participants with severe or function-limiting pain without emergency warning signs were referred for same-day urgent-care evaluation. Emergency-department or 911 referral was reserved for emergency warning signs, major trauma, sudden or rapidly worsening severe pain, or another time-sensitive medical concern. Further referrals included the distribution of information for free phone programs to ensure better healthcare communication for participants without reliable telephone access.

Any eligible encounters during the predefined implementation period were included in the data analysis. No inferential power calculation was performed because the project was designed as a descriptive operation improvement initiative and not a hypothesis testing study. As this implementation project was not designed to evaluate causal effects of the outreach implementation, the study cannot determine whether the pathway reduced emergency department utilization, improved pain, increased completed primary-care visits or impacted long-term health outcomes.

Contact information was collected from participants who consented by providing their telephone numbers to Valliant Foundation Homeless heartline, which was kept in a separate file, unlinked to the quality-improvement questionnaire. Scheduled follow-up calls were offered to participants to provide non-clinical peer support, supportive listening and assistance with navigating community resources, including healthcare, housing, food assistance and other social services available; however, these services were not provided as a substitute for medical, mental health or emergency services.

### 10. Measures

#### Program Development and Volunteer Training

Volunteers completed standardized classroom and field training developed by medical professionals and reviewed by the Ethics Committee. Training covered questionnaire administration, pain assessment, vital-sign measurement, equipment use, documentation, first aid, blood-borne pathogens, recognition of abnormal findings, and emergency escalation. Volunteers shadowed outreach officers and demonstrated competency by performing required tasks under supervision before independently participating in screening.

An outreach officer was required to witness correct demonstration of vital sign collection and the completion of assessments on outreach practices with 100% accuracy by the volunteer to ensure proper understanding of equipment and protocols. Before using the glucometer, outreach volunteers were required to complete at least ten supervised demonstrations before they could independently measure and capture glucometer data and capillary specimen collection. In the study, no assessment of inter-rater reliability was made using an intraclass correlation coefficient. Repeated measurement on a single participant by three volunteers was deemed ethically inappropriate and impractical in this QI outreach protocol based on the vulnerable, transient population. Instead, accuracy and consistency of measurements were supported by training sessions, demonstrated proficiency, and immediate reviews of abnormal data by supervising health officers. Through this approach, participant dignity was prioritized while data quality was maintained by immediate oversight.

**Table 1.** Participant characteristics and healthcare access.

| *Characteristic* | *Valid n* | *n (%) or summary statistic* |
| --- | --- | --- |
| **Age, years** | 184 | 43.36 ± 10.96; median 41.5 (22–72) |
| **Sex** | 193 |  |
| Male | 193 | 139 (72.0) |
| Female | 193 | 44 (22.8) |
| Unknown or missing | 193 | 10 (5.2) |
| **Race/ethnicity** | 193 |  |
| White | 193 | 70 (36.3) |
| Black or African American | 193 | 63 (32.6) |
| Hispanic or Latino | 193 | 18 (9.3) |
| American Indian, Alaska Native, or<br>Indigenous | 193 | 7 (3.6) |
| Other or specified origin | 193 | 22 (11.4) |
| Unknown or missing | 193 | 13 (6.7) |
| **Reports access to a PCP or specialist** | 115 |  |
| Yes | 115 | 55 (47.8) |
| No | 115 | 60 (52.2) |

\* Summary of extracted methodology and the core values retrieved for the emergency response protocol that represents standard protocol guidelines.

Participation was limited to adults experiencing homelessness who demonstrated adequate decision-making capacity and voluntarily provided informed consent. Capacity was assessed pragmatically using orientation questions and teach-back to confirm understanding of the assessment before participation.

Exclusion criteria included the absence of decisional capacity due to altered mental status, visible signs of drug or alcohol intoxication, or withdrawal of consent at any stage of participation. To ensure voluntary participation and participant safety, outreach teams were trained to assess decisional capacity before proceeding with study activities. If a participant demonstrated signs of disorientation or impaired capacity, the assessment was immediately terminated in accordance with the “Hard Stop” protocol. Inclusion/exclusion criteria are outlined in Figure 1.

**Figure 1.**
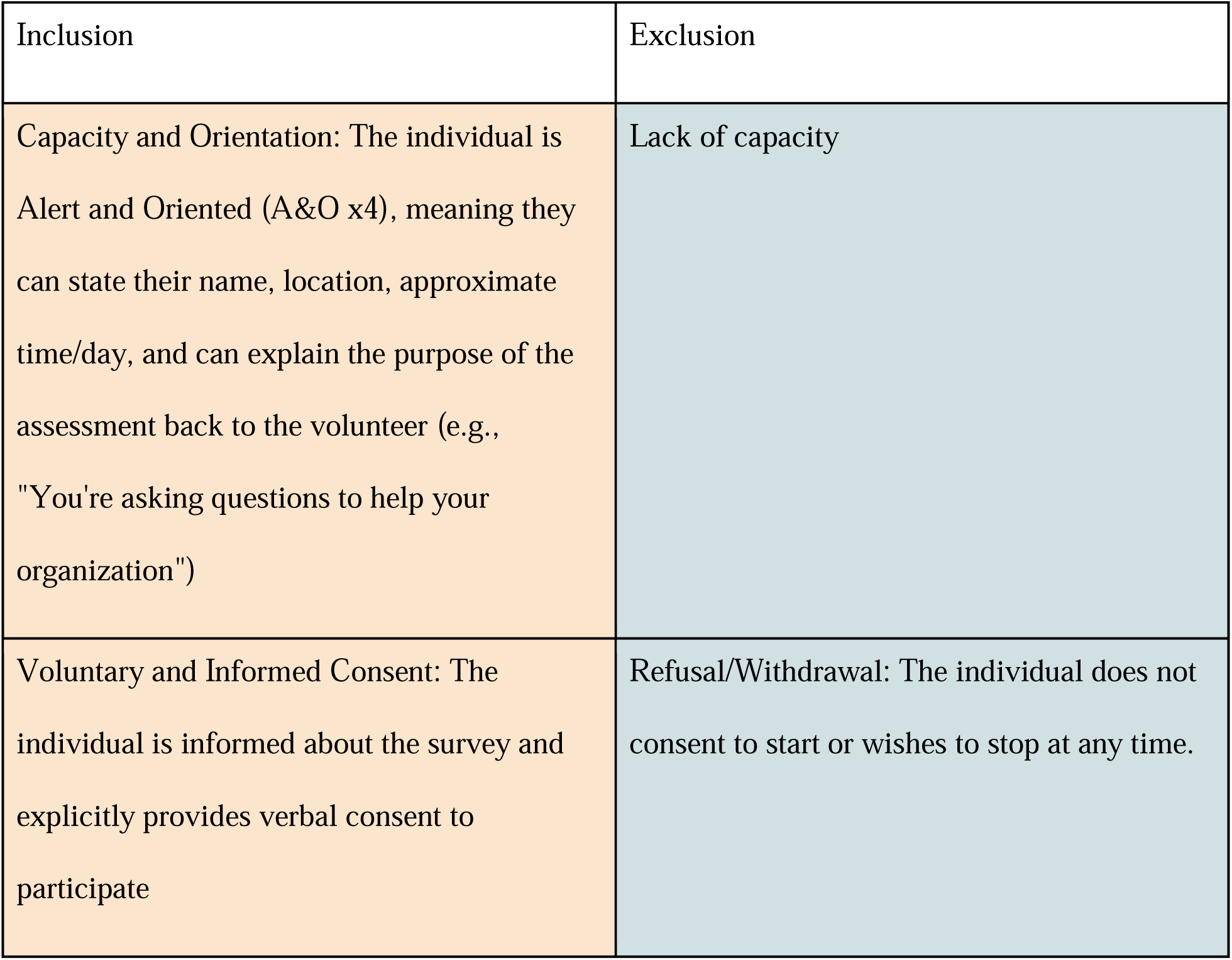
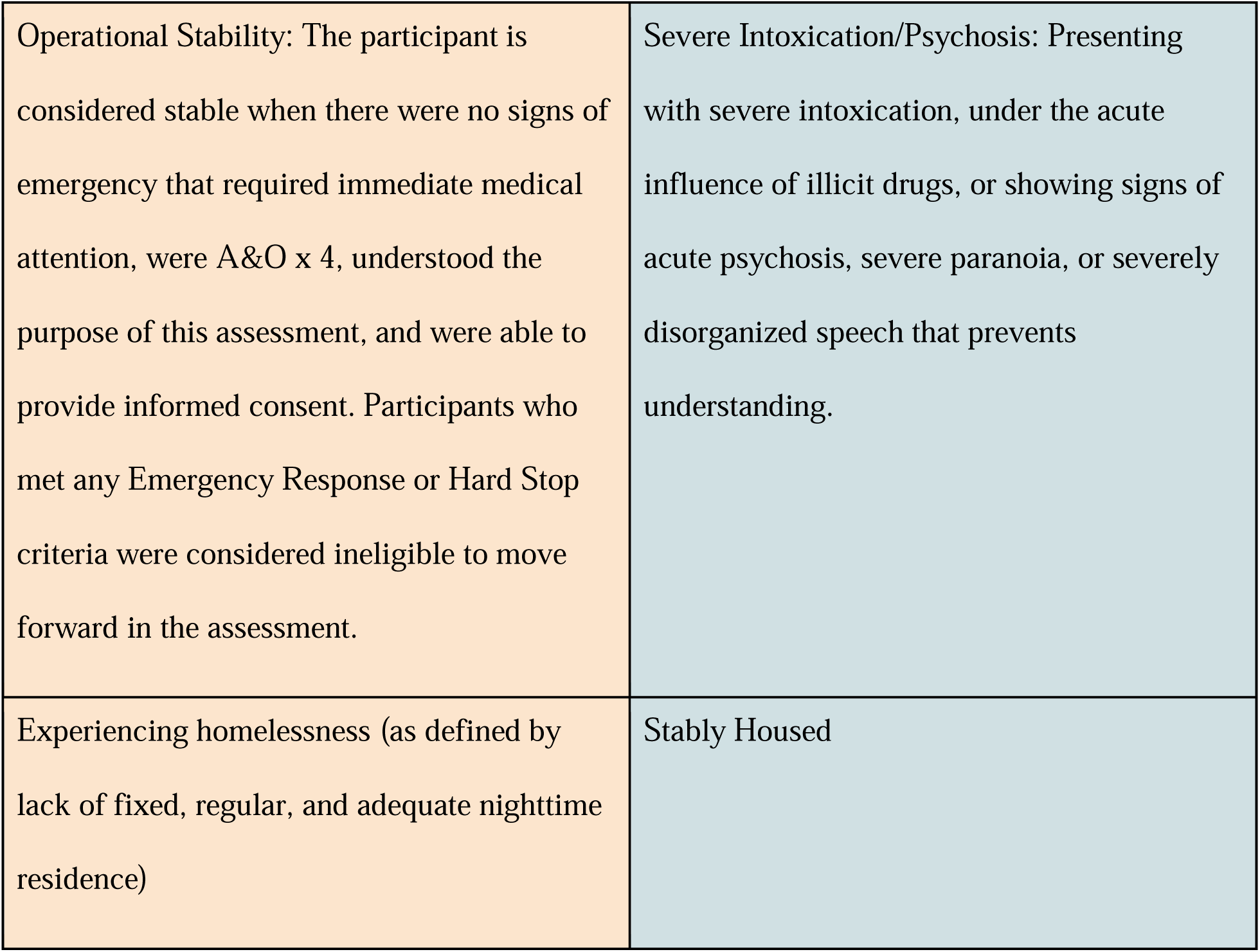
Flow of Participant Screening, Inclusion, and Exclusion Based on Eligibility Criteria.

#### Verbal Consent

Volunteers explained the purpose of the assessment, emphasized that participation was voluntary, and obtained verbal informed consent before beginning any screening activities.

Participants could decline or withdraw at any time, at which point the assessment ended immediately according to the established Hard Stop procedure.

#### Data Collection

The questionnaire collected demographic and health-related information, including age, gender, race/ethnicity, substance and alcohol use, diabetes status, and nicotine use. In addition, two physiological measures (blood pressure and heart rate) and one subjective measure (self-reported pain score) were obtained.

Data were collected across the Inner Mission, Tenderloin, and Embarcadero neighborhoods of San Francisco and Sacramento, California. Outreach team members approached participants and, if the inclusion criteria were met and informed consent was provided, administered the questionnaire and collected relevant vital-sign data. This convenience sampling approach may introduce selection bias, as individuals who choose to interact with outreach teams may be better connected to community resources or may have sought engagement due to awareness of pre-existing cardiovascular conditions (self-selection bias). To avoid duplicate data collection from the same sources, the protocol used verbal screening to confirm prior participation before any data were recorded. This approach ensured that the dataset remained cross-sectional rather than longitudinal.

Data were collected through convenience sampling during routine outreach. Eligible participants completed an orally administered questionnaire and field health assessment. Physiological measurements were obtained using portable medical devices according to standardized field procedures and under trained supervision. Equipment was inspected before use, and irregular measurements were repeated or reviewed when feasible. Because data collection occurred in nonclinical environments, standard procedures were adapted when necessary to preserve participant safety, dignity, and engagement.

In addition to physiological measures, subjective pain intensity was assessed using the Numeric Pain Rating Scale (NPRS) [9], a standardized instrument for quantifying self-reported pain. Participants were asked to rate their current pain on a scale of 1 to 10. Consistent with San Francisco Emergency Medical Services (EMS) practice, which applies the NPRS on a 1–10 range, responses of “0” were recorded to “1” and values exceeding “10” were recorded to “10.” This transformation reflects alignment with local clinical standards rather than a modification of the underlying pain construct. Pain score data were collected for descriptive and operational purposes only and were not used for diagnostic inference.

Narrative observations were collected to provide contextual illustrations of engagement rather than the formal outcomes of the measures, and should not be interpreted as evidence of clinical or psychosocial impact. Follow-up measures, such as clinic attendance, consultations, and blood pressure control, were beyond the scope of this methodology. As an alternative, all participants were offered a handout listing local shelters and free health clinics. Participants with access to a cell phone were offered information about the volunteer-run hotline Homeless Heartline for follow-up.

All data were collected anonymously in the field, with each participant assigned a random string of letters and numbers as an identifier; no direct identifiers [8], including, but not limited to, names or Social Security numbers, were collected. Phone numbers were collected in a separate service record that is not linked to the evaluation dataset. Furthermore, a comprehensive effort was made to avoid groups of indirect identifiers that could risk re-identification, including all elements of dates related to the individual (except the year), precise location, and photographs or biometric data. Data analysis for reports also clustered sensitive information: for example, all diabetes types were grouped to reduce the likelihood of identifying an individual with a rarer type, such as Type 1.

To maintain security, data files are stored in password-protected files, accessible only by the Principal Investigator and a select few authorized project managers. Regular monitoring of collected data immediately after each collection event must be performed to screen for direct or quasi-identifiers to reduce the likelihood of possible re-identification. If it was discovered that a direct identifier or quasi-identifier was collected by mistake, the entire data item for that participant was immediately deleted. In line with institutional best practices, once data collection was complete, these files were encrypted. The documents will be retained for up to 5 years before being automatically deleted from institutional servers, thereby ensuring the long-term protection of participants while supporting the project’s aim to provide a safe, transparent, and reproducible model for other organizations.

#### Emergency Response Protocol

Participants demonstrating emergency warning signs were offered 911 activation according to predefined escalation criteria. Participants retained the right to decline assistance if they had decision making capacity as defined as alert and oriented times four (A&Ox4) and lacked suicidal and homicidal ideation. Abnormal vital signs, chest pain, signs of stroke, or recent major trauma are examples of events that would trigger the Emergency Response Protocol.

### 11. Analysis

A descriptive analysis of field notes was employed to evaluate the implementation of the Mobile Outreach Initiative (MOI) over a 19-month period. Field observations and volunteer notes were iteratively reviewed to identify recurring themes related to participant autonomy, barriers to care, and the performance of the Emergency Response Protocol. Qualitative data were analyzed within a structured framework encompassing protocol adherence, longitudinal engagement, and informed consent (see Table 2).

**Table 2.** Pain and self-reported emergency-department utilization.

| *Measure* | *Valid n* | *n (%) or summary statistic* |
| --- | --- | --- |
| **Current pain score** | 175 |  |
| Mean ± SD | 175 | 3.79 ± 2.78 |
| Median (range) | 175 | 3 (1–10) |
| **Self-reported ED visits during the specified recall period** | 99 |  |
| 0 visits | 99 | 43 (43.4) |
| 1 visit | 99 | 22 (22.2) |
| 2–3 visits | 99 | 19 (19.2) |
| 4–5 visits | 99 | 7 (7.1) |
| 6 or more visits | 99 | 8 (8.1) |

All participants (N=193) were screened using an A&O x4 assessment to confirm cognitive status before proceeding with data collection. Because mental status is a prerequisite for valid consent, all participants included in the study were screened with the A&O x4 assessment.

### 12. Ethical Considerations

Survey instruments were standardized, and participant data anonymized; interactions were also standardized across pre-survey, survey, and post-survey phases. Prior to commencement, the project was reviewed by Valliant Foundation’s *Ethics Committee*, an independent oversight body authorized by the Foundation’s bylaws to review, approve, and enforce ethical standards for operational and research activities.

The Committee determined that this project constitutes Non-Human Subjects Research (NHSR) under 45 CFR 46.102, as it does not aim to contribute to generalizable knowledge and was conducted solely to inform nonprofit operations and outreach programming. Regular check-ins with the committee were performed to maintain ethical oversight and monitoring throughout the project.

#### Field Implementation

To reduce the potential for coercion and ensure ethical engagement, volunteers first provided access to food and essential resources before discussing opportunities to participate in surveys. This approach prioritized participants’ immediate well-being and mitigated undue influence in populations experiencing resource scarcity. Before conducting any physiological assessment, volunteers performed a cognitive capacity screening to confirm participants’ ability to provide informed consent and proceeded only after obtaining verbal consent. All volunteers received structured training in standardized physiological data collection procedures and culturally responsive health education, aligned with program standards and ethical best practices.

The total count of individuals engaged during the greeting phase was not formally tracked, as the focus remained on immediate outreach. Notably, some individuals declined the full screening, but still engaged with the team for a brief physical check-up and to receive essential supplies, such as food and water. Regardless of the refusal, volunteers immediately provided food and water before requesting consent to participate in the survey.

### 13. Results

Pain scores were available for 175 participants (valid n = 175). Category percentages are based on 165 categorized pain scores because some values were uninterpretable. The mean pain score was 3.79, SD = 2.78, the median was 3, and the range was 1–10. Based on the modified Numerical Rating Scale used in this study, pain scores were categorized as no pain (1), mild (2– 3), moderate (4–6), and severe (7–10). A total of 53 (32.1%) participants reported no pain, 29(17.6%) reported mild pain, 49(29.7%) reported moderate pain, and 34 (20.6%) reported severe pain.

The Mobile Outreach Initiative (MOI) protocol built upon an existing ethically reviewed outreach framework that incorporated participant capacity screening, verbal informed consent, the Hard Stop procedure, standardized volunteer training and oversight, vital sign and pain assessment, emergency escalation and resource navigation. These components established a safe and standardized foundation for community-based outreach. However, although pain was routinely identified, volunteers lacked a standardized process for responding after pain was recognized, resulting in variability in referral decisions and supportive care.

The intervention introduced a standardized pain assessment, a tiered referral pathway, optional hot and cold packs, and expanded resource navigation. Standardized assessment was intended to reduce volunteer uncertainty, while the tiered pathway improved consistency in directing participants to the appropriate level of care. The pathway was designed to direct nonemergent pain toward primary or urgent care while preserving emergency escalation for red-flag findings and reducing inappropriate default referral to emergency departments. Hot and cold packs provided immediate, low cost, portable, nonpharmacologic comfort that was compatible with the screen and refer model and respected participant autonomy. Expanded resource navigation addressed barriers such as limited access to telephones, transportation, insurance, clinic information and stable housing that could prevent participants from acting on referral recommendations. Consistent with quality-improvement methodology, community engagement informed refinement of the protocol rather than hypothesis generation or study governance.

**Table 3.**
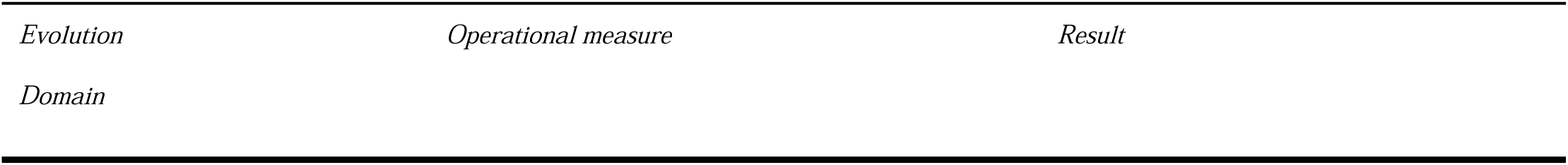
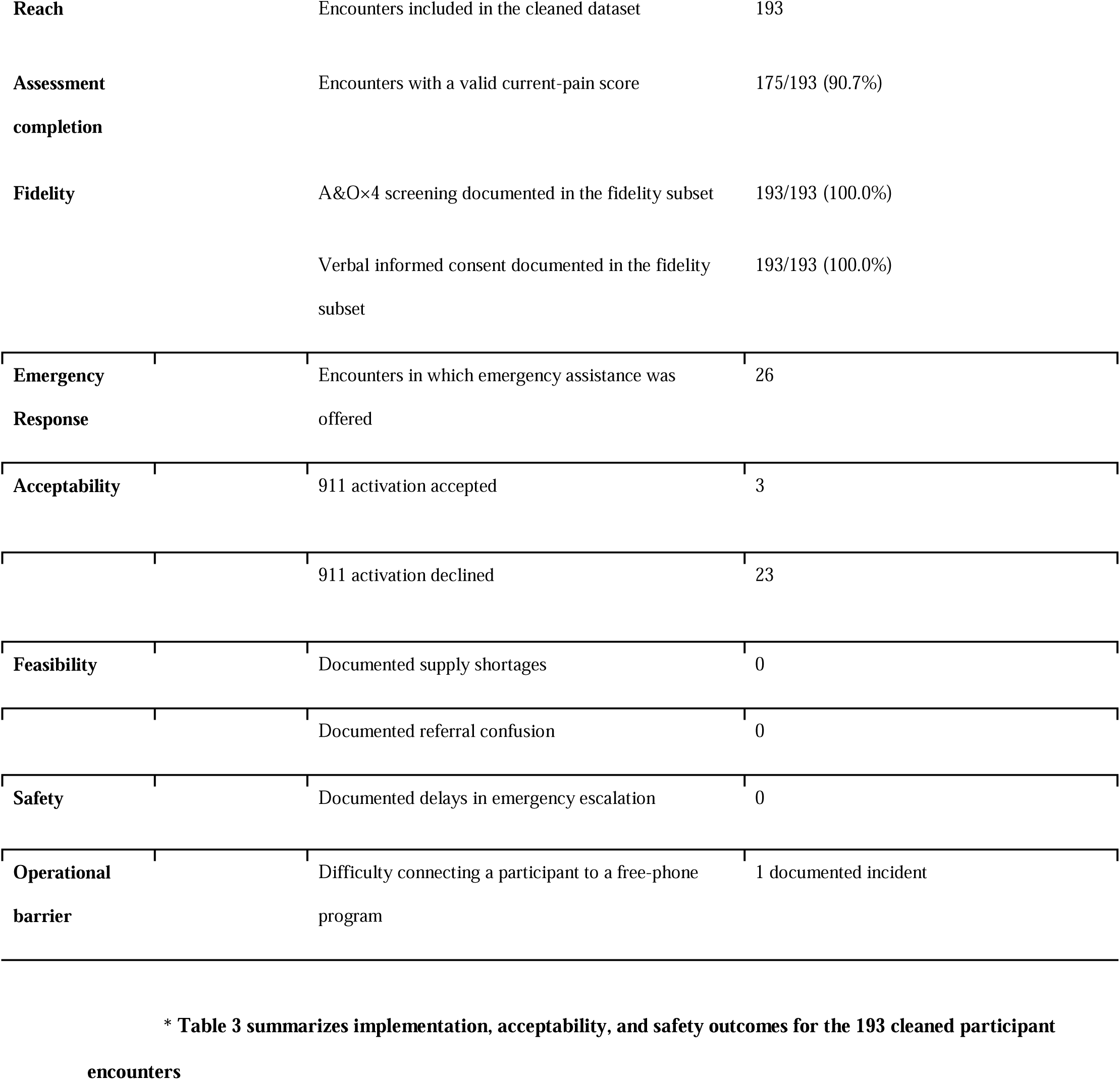
Implementation, acceptability, and safety outcomes.

**Table 4.** Tiered pain-response criteria.

| *Metric* | *Sample Size (N)* | *Mean* |
| --- | --- | --- |
| **Emergency pain** | Pain with emergency warning signs, major trauma, sudden severe onset, or rapid worsening | Offer immediate 911 activation or emergency-department evaluation |
| **Severe or function-limiting pain** | Pain rated 7–10 or substantially limiting movement or daily activities, without emergency warning signs | Refer for same-day urgent-care evaluation |
| **Stable nonemergent pain** | Stable mild, moderate, or chronic pain without emergency warning signs | Refer to primary care or an appropriate community clinic |
| **Cold-pack indications** | Recent minor injury, sprain, strain, swelling, bruising, overuse, or localized acute musculoskeletal pain | Offer an optional cold pack for 15–20 minutes with a skin barrier |
| **Hot-pack indications** | Muscle tightness, cramps, spasms, stiffness, arthritis-related stiffness, or stable chronic musculoskeletal pain | Offer an optional hot pack for 15–20 minutes with a skin barrier |
| **Pack contraindications** | Broken skin, active bleeding, impaired sensation, suspected infection, fracture, serious injury, or possible delay of emergency care | Do not provide a pack; refer to the appropriate level of care |

\*Table 4 summarizes the tiered pain-response criteria implemented during the outreach protocol.

**Table 5.** Self-reported clinical conditions relevant to pain assessment.

| Condition group | Valid n | Condition present, n (%) |
| --- | --- | --- |
| Behavioral-health or psychiatric condition | 120 | 73 (60.8) |
| Substance-use or addiction-treatment context | 120 | 57 (47.5) |
| Musculoskeletal, orthopedic, or injury-related condition | 120 | 43 (36.1) |
| Cardiovascular or vascular condition | 120 | 39 (32.5) |
| Neurologic, neurotrauma, or neuropathic-pain risk | 120 | 32 (26.7) |
| Wound, skin, or soft-tissue condition | 120 | 24 (20.0) |
| Respiratory or pulmonary condition | 120 | 19 (15.8) |
| Endocrine or metabolic condition | 120 | 13 (10.8) |
| Infectious disease or bloodborne virus | 120 | 9 (7.5) |
| Cancer, immune/rheumatologic, pregnancy, or other high-complexity flag | 120 | 2 (1.7) |

**Table 5 reports the frequency of self-reported clinical conditions relevant to pain assessment. Percentages are based on valid responses.**

Healthcare access remained limited among participants. Of the 115 participants with valid responses, 55 (47.8%) reported access to a primary care provider or specialist, whereas 60 (52.2%) reported no such access. Among the 99 participants with valid responses regarding emergency department use during the previous year, 43 (43.4%) reported no visits, while 56 (56.6%) reported one or more visits. Because access to a primary care provider and specialist was collected as a single combined variable, these findings cannot distinguish primary-care access from specialist access alone.

**Table 6.** Operational Themes.

| *Theme* | *Field Evidence* | *Evaluation or planning implementation* |
| --- | --- | --- |
| Exertion-related pain and mobility burden | “Heel and ankle pain during prolonged periods of walking” | Functional and environmental context should accompany the numerical pain score |
| Persistent injury-related pain | “5/10 to left ankle after traumatic injury that occurred within the last year” | Stable trauma-related pain may require longitudinal primary-care or specialty follow-up |
| Severe localized pain | “8/10 pain that remains at the site” | Severe pain requires same-day evaluation even when emergency warning signs are absent |
| Limited telephone access | Participant “currently does not have a phone but wanted medical assistance” | Referral completion may depend on communication assistance, not merely providing clinic information |
| Comfort-measure refusal | A hot-pack offer for elbow and knee pain “was rejected” | Acceptability should be measured separately from whether the intervention was available |
| Difficulty connecting to resources | Volunteers documented difficulty enrolling a participant in a phone program | Resource-navigation feasibility should include unsuccessful connection attempts |

Several recurring themes related to pain management and referrals were identified during outreach. Participants with chronic illnesses, previous injuries, or pain that limited their mobility often had difficulty traveling to clinics or following through with referrals. Limited access to phones, internet service, and transportation created additional barriers to follow-up care. Some participants declined referrals because of previous negative experiences with healthcare, concerns about stigma, transportation challenges, or competing priorities such as obtaining food or shelter. Volunteers also reported documentation challenges, including equipment malfunctions, interrupted interviews, incomplete medical histories, and unreliable internet connectivity. An additional challenge arose during data analysis due to missing or unknown data.

Initial missions encountered difficulties maintaining device calibration and ensuring privacy in busy public spaces. To address this, teams added a second blood pressure monitor for cross-verification and implemented a “quiet zone” policy.

#### Details about Missing Data

Missing pain scores occurred in 18 participants (9.33%). Pain score missingness may reflect both Missing at Random (MAR) and Missing Not at Random (MNAR) patterns. MAR may have resulted from interrupted outreach encounters or incomplete documentation during field-based outreach, whereas MNAR may have occurred when participants experiencing severe pain were unable or unwilling to complete the assessment. Referral tier and hot/cold pack disposition were not routinely documented because recording every intervention during time-limited outreach encounters would have substantially increased the documentation burden. Healthcare access and emergency department utilization data may also have been missing because some participants needed emergency assistance or too ill to complete the assessment, and others may have been reluctant to disclose their access to primary or emergency care. Additional implementation barriers included limited clinic availability and lack of reliable telephone access among a substantial minority of participants. No duplicate records or supply shortages were identified during the implementation period.

**Table 7.** Data Integrity Summary: Rationale for Outlier Exclusion and Missing Data.

addresses the “Missing at Random” pattern primarily stems from logistical barriers inherent to mobile outreach and data were also excluded if values were determined to be physiologically implausible.
| *Issue* | *Frequency (N)* | *Impact on Analysis* | *Resolution* |
| --- | --- | --- | --- |
| Physiologically Implausible BP | N= 1 | Excluded from BP mean/range. | Manual review for accuracy; data points like 135/32 were deleted due to logistical barriers. |
| Extreme HR Outliers | N= 1 | Excluded from HR mean/range. | HR readings such as 825 bpm were removed to preserve analytical integrity. |
| Missing/non-analyzable pain data | N=18 | Excluded from pain mean/range |  |
| Accidental Identifiers | N= 0 | Item-level detection. | Per protocol, any entry containing a name or phone number was immediately deleted. |
| Withdrawal of Consent (Refusal) | N/A | Immediate cessation. | "Hard Stop" Protocol: Assessments were discontinued if impairment was observed, capacity was lost, or refusal occurred. |

Increasing the sample size would have been difficult given the Foundation’s scope of resources. Moreover, we acknowledge this as a constraint, as it limits our ability to quantify the direction or magnitude of selection bias or to calculate a formal participation rate.

## Discussion

### 14. Summary

This operational needs assessment yielded measurable implementation outcomes and practical insights into field-based pain assessment and referral among adults experiencing unsheltered homelessness. The final dataset included 193 encounters, with valid pain scores available for 175 participants; mean pain intensity was 3.79 ± 2.78, with a median score of 3. Pain scores ranged from 1 (no pain) to 10 (severe pain) utilizing the Numeric Pain Rating Scale. The intervention translated pain scores and emergency warning signs into standardized recommendations for primary care, same-day urgent care, or emergency services. Cold packs were used for selected acute discomfort and foot or ankle pain, while hot packs were used for joint pain; one participant declined an offered comfort measure. Emergency assistance was offered in twenty-six encounters and accepted in three. Among the 193 encounters included in the implementation-fidelity assessment, participant capacity screening and informed verbal consent were documented in all cases. No supply shortages, referral confusion, or delays in emergency escalation were documented, although limited telephone access remained an important barrier to completing follow-up care.

Relative to existing outreach models, this framework occupies a distinct implementation niche. Unlike resource-intensive mobile clinics that emphasize diagnostic capacity, treatment, and longitudinal management, this model prioritizes rapid, low-resource, field-based pain screening, ethical engagement, immediate nonpharmacologic comfort measures, and referral to the appropriate level of care in transient environments. In contrast to Community-Based Participatory Research approaches, which emphasize shared governance and hypothesis generation, this initiative was operationally focused and emphasized immediate service delivery, standardized decision-making, and iterative protocol refinement. Although implementation was feasible within the participating outreach settings, scalability, referral completion, pain outcomes, and broader applicability require further evaluation using formal implementation and longitudinal outcome measures.

### 15. Interpretation

A central contribution of this initiative was demonstrating that standardized pain-related decision-making could be incorporated into low-resource street outreach while preserving participant autonomy and emergency safeguards. The pathway gave volunteers a consistent method for distinguishing emergency conditions from stable nonemergent pain while allowing temporary comfort measures when appropriate.

Referral recommendations did not eliminate structural barriers to care. Limited telephone and transportation access, difficulty obtaining appointments or medications, financial constraints, institutional mistrust, clinic eligibility requirements, and participant mobility continued to affect follow-through [3,18]. Participant refusal of recommended emergency assistance further demonstrated that identifying the appropriate level of care does not guarantee that the service will be accessible or acceptable. These findings support the use of low-resource, trust-centered outreach models that combine standardized pain triage with individualized resource navigation.

Overall, the observed outcomes were largely consistent with the initial expectations. The primary deviations from expectations being that supply shortages were anticipated during outreach, but no shortages of assessment materials or comfort measures were observed. Furthermore, while greater Emergency Medical Service activation was expected, participants declined emergency assistance due to cost considerations and hesitancy toward receiving unprejudiced treatment during ED visits. These findings suggest that barriers to receiving emergency care were related to prior negative experiences rather than limitations of the referral pathway.

### 16. Limitations

This study has several limitations inherent to its design as a quality-improvement operational needs assessment. Convenience sampling without a formal sampling frame introduces potential selection bias because individuals who engaged with outreach teams may have differed systematically from those who declined participation, lacked decision-making capacity, were acutely intoxicated, or were not present during scheduled outreach. The total number of individuals approached was not recorded; therefore, participation rates could not be calculated. Refusal data were also incomplete. Emergency assistance was offered during twenty-six encounters and accepted during three, resulting in six documented refusals, but refusals of the questionnaire, individual physiological measurements, pain assessment, and resource navigation were not consistently recorded. These limitations prevent estimation of the direction or magnitude of selection bias.

The study was geographically, environmentally, and temporally constrained. Outreach was conducted in selected urban neighborhoods in San Francisco and Sacramento, primarily during daylight hours and favorable weather conditions. Individuals who were more accessible at night, unable to travel to the outreach locations, experiencing severe mobility limitations, or sheltering from adverse weather may therefore be underrepresented. The protocol also excluded individuals who could not complete the field-based orientation and consent process. Although this protected participants who could not safely provide consent, it likely excluded individuals experiencing severe intoxication, acute psychosis, cognitive impairment, or other conditions associated with particularly high medical and pain-related needs. The findings should consequently be interpreted as context-specific observations from individuals able and willing to participate in daytime outreach rather than population estimates for all people experiencing unsheltered homelessness.

Pain measurement was based primarily on a single self-reported numerical rating of current pain. Although the Numeric Pain Rating Scale offers a practical field measure, a point-in-time score may not capture intermittent, recurrent, or function-limiting pain. Narrative responses demonstrated that some participants reported little or no pain during the encounter despite describing usual pain of greater intensity, chronic stiffness, or episodic back pain severe enough to prevent walking. Other participants described pain that intensified with exertion, prolonged walking, stress, or changing circumstances. The numerical score therefore should not be interpreted as a comprehensive measure of longitudinal pain burden, functional impairment, or treatment need. Self-reported diagnoses, medication use, injury history, and associated symptoms were not independently verified through physical examination or medical records.

The qualitative findings are also limited by the nature of the source material. Narrative data consisted of brief participant responses and volunteer-entered field notes rather than recorded, transcribed qualitative interviews. The amount of detail varied substantially across encounters, and some entries were likely paraphrases rather than verbatim statements. Environmental noise, time constraints, participant fatigue, intoxication, communication difficulties, and differences in volunteer documentation practices may have affected the completeness and accuracy of the narratives. The thematic findings should therefore be considered descriptive and illustrative rather than a comprehensive qualitative account of the lived experience of pain.

Primary-care access and emergency-department utilization were self-reported and may be affected by recall error, uncertainty regarding the type of facility visited, and difficulty remembering the number, timing, or reason for prior encounters. Medical records were not available to verify reported diagnoses, ED visits, hospitalizations, medication prescriptions, or primary-care appointments. Some participants listed multiple reasons for emergency care, including pain, trauma, overdose, infection, and mental-health crises, making it difficult to classify a visit as exclusively pain-related. Similarly, reporting contact with a primary-care clinician did not establish timely access, continuity with the same clinician, completion of specialty referrals, or successful medication management. Narrative responses indicated that some participants technically had a primary-care provider but still experienced difficulty obtaining appointments or refills, while another reported more than ten ED visits despite recent primary-care contact.

The pain-specific intervention was implemented during a limited portion of the overall outreach period. Hot and cold packs were available only during the designated implementation period, while participants encountered earlier received the preexisting outreach protocol without these comfort measures. The limited intervention exposure prevents reliable estimation of acceptability across the broader population and does not permit conclusions regarding the effectiveness of hot or cold packs in reducing pain. Documentation also did not consistently capture pain ratings after pack use, duration of relief, adverse skin effects, or whether participants followed the recommended referral.

The iterative development of the protocol represents an important quality-improvement strength but complicates interpretation. The outreach framework evolved over time in response to device problems, privacy concerns, incomplete documentation, encounters with vulnerable participants, and the identification of a pain-management gap. Participants were therefore exposed to different versions of the protocol depending on when they were encountered. Without a contemporaneous comparison group, a standardized preimplementation period, or formal pre– post analyses, observed implementation outcomes cannot be attributed solely to the final pain-response pathway. The study was not designed to determine whether the intervention reduced pain, increased primary-care attendance, improved medication access, or decreased subsequent emergency-department utilization.

Missing, incomplete, and excluded data introduce additional uncertainty. Variable-specific denominators differed because not every participant provided complete information and some physiological values were removed after being judged implausible. Complete-case analyses may be biased if participants with missing pain, utilization, or access data differed systematically from those with complete responses. Missingness may have resulted from logistical and documentation problems consistent with Missing at Random mechanisms, but Missing Not at Random mechanisms are also plausible when participants declined measurements because of anxiety, pain, mistrust, intoxication, or concern about abnormal findings. Sensitivity analyses were not performed, limiting evaluation of how alternative assumptions about missing data would affect the findings.

Although volunteers completed structured training and worked under supervision, inter-rater reliability was not formally evaluated. Differences in experience may have influenced the identification of emergency warning signs, interpretation of functional impairment, selection of hot or cold packs, and documentation of qualitative responses. The A&O×4 process was used as a pragmatic situational-safety and orientation screen rather than a validated assessment of decisional capacity. It should therefore not be interpreted as a diagnostic cognitive evaluation or proof of formal capacity. Standardized training, competency demonstrations, supervisory consultation, and predefined referral criteria reduced, but could not eliminate, variation between volunteers.

Finally, the absence of longitudinal follow-up restricts interpretation to immediate implementation performance. The study did not systematically determine whether participants attended recommended primary-care, urgent-care, or emergency services; obtained medications; completed postoperative or specialty follow-up; experienced subsequent changes in pain; or returned to the emergency department. Telephone loss, unstable contact information, transportation barriers, and population mobility limited the feasibility of follow-up and may also be central mechanisms through which referrals fail. Future evaluations should incorporate referral-tracking procedures, repeated pain and functional assessments, participant-reported experience measures, verification of healthcare utilization when ethically and operationally feasible, and implementation across more diverse geographic and service settings.

Despite these limitations, the standardized procedures for capacity screening, verbal consent, emergency escalation, pain assessment, tiered care recommendations, comfort measures, and resource navigation support procedural reproducibility. The findings demonstrate that a structured pain-response pathway can be incorporated into low-resource outreach operations, but they should not be interpreted as evidence of clinical effectiveness or reduced emergency-department utilization. Broader scalability and impact require prospective evaluation using clearly defined implementation measures, longitudinal outcomes, and comparison across different outreach organizations and populations.

#### Lessons Learned

Operational challenges and iterative adaptations improved subsequent Mobile Outreach Initiative missions and clarified several priorities for future evaluation and program planning. The Emergency Response Protocol functioned effectively as a living document, but future implementations should prospectively define the baseline protocol, the date and rationale for each modification, and the specific outcomes expected from each change. Maintaining a version-controlled protocol log would allow evaluators to distinguish outcomes generated under earlier procedures from those observed after implementation of the tiered pain-referral pathway, hot-and cold-pack interventions, and expanded resource navigation. Protocol revisions should also be communicated through structured update sessions, written change summaries, and brief competency checks so that all volunteers apply the current procedures consistently.

Volunteer preparation should address both technical skills and the interpersonal and operational challenges of street-based outreach. In addition to pain screening, vital signs, and referral thresholds, training should incorporate cultural humility, recognition of personal bias, trauma-informed communication, active listening, de-escalation, and scenario-based practice involving emergency-service refusal, intoxication, mental-health crises, and severe nonemergent pain. Programs should also anticipate field barriers such as limited privacy, environmental noise, participant mobility, and portable-equipment limitations by establishing quieter assessment areas, backup equipment, procedures for confirming implausible measurements, and consultation for red-flag findings before implementation. Refresher training should accompany changes to referral criteria or safety procedures so that these safeguards are incorporated prospectively rather than added in response to field challenges.

For many volunteers, participation in the Mobile Outreach Initiative represented an initial experience with field-based public-health service. Although this created opportunities to develop patience, empathy, cultural humility, active listening, and de-escalation skills, volunteer learning was not initially measured as a formal implementation outcome. Future evaluations should assess volunteer preparedness before and after training, document protocol deviations, and obtain structured feedback regarding usability, decision-making confidence, and barriers encountered during implementation. These measures would help determine whether training improves fidelity and would identify areas requiring additional supervision or protocol simplification.

Data quality and referral follow-through should be planned prospectively. Early implementation was affected by incomplete, missing, and erroneous entries, while the absence of systematic longitudinal follow-up limited assessment of whether participants completed primary-care, urgent-care, or emergency-department referrals. Future programs should use real-time data review, required fields, standardized definitions, and end-of-mission record reconciliation to identify errors while encounters remain recent. Evaluators should also record the number of individuals approached, survey and measurement refusals, referrals offered, referrals accepted or declined, comfort measures offered, and barriers to follow-up. When ethically and operationally feasible, referral tracking should include repeated contact attempts, appointment completion, subsequent pain status, and emergency-department utilization. These changes would strengthen interpretation of feasibility, fidelity, acceptability, and program reach while preserving the flexibility required for low-resource street outreach.

### 17. Conclusion

This project demonstrates that ethical safeguards can be integrated into mobile health outreach alongside a standardized pain-response pathway. The framework combined capacity screening, verbal consent, the “Hard Stop” procedure, pain assessment, tiered care recommendations, and predefined emergency escalation.

Implementation outcomes included feasibility, care-level recommendations, participant acceptance or refusal of comfort measures, emergency response, and operational barriers. Hot and cold packs were added as optional, temporary comfort measures, while documentation and telephone-access challenges informed continued protocol refinement.

These findings suggest that structured pain assessment, referral guidance, and nonpharmacologic support can be incorporated into outreach while preserving participant autonomy. The initiative was not designed to determine whether pain improved, referrals were completed, or emergency-department use decreased.

This framework provides a replicable, low-resource model for organizations seeking to strengthen pain assessment, referral practices, ethical standards, and access to appropriate healthcare for unsheltered populations.

## Data Availability

All data produced in the present study are available upon reasonable request to the authors

## 18. Other Information

### Ethics Statement

This project was reviewed by Valliant Foundation Ethics Committee and determined to constitute Non-Human Subjects Research under 45 CFR 46.102.

### Declaration of generative AI and AI-assisted technologies in the manuscript preparation process

During the preparation of this work, the authors used NotebookLM to improve grammatical, spelling, and organizational clarity. After using this service, the authors reviewed and edited the content as needed and took full responsibility for the published article.

### Author Contributions

Redacted

## Appendix A: Definitions

**Capacity Screening (A&O x4)**-volunteers must verify the participant is Alert and Oriented (A&O x4). The individual must be able to state: (1) Full name, (2) Current location, (3) Approximate time/day, and (4) Purpose of the assessment (e.g., “You are checking my health to help your organization”).

**Consent Procedures**-verbal consent from all participants is required before any physiological measurements can take place. Participants are informed of their right to withdraw at any time, for any reason, without penalty or loss of services. Participation is non-coercive, strictly voluntary, and independent of resource distribution. Staff were required to emphasize that participation was strictly voluntary and entirely independent of the distribution of humanitarian resources. Ethical integrity was maintained by ensuring that refusal or withdrawal from the screening carried no penalty and did not impact the participant’s eligibility for other outreach services.

**Emergency Response Protocol**-a stepwise procedure outlining actions for volunteers to take when encountering participants with urgent or life-threatening vital signs or physiological abnormalities.

**Exclusion Criteria**-Individuals who were stably housed or otherwise failed to meet the definition of unsheltered homelessness were ineligible for participation. Individuals were excluded if they lacked the cognitive capacity to provide informed consent, including those exhibiting disorientation, acute psychosis, or severe paranoia. Individuals who were under the apparent influence of illicit drugs or alcohol to a degree that prevented an informed understanding of the screening process were also excluded.

**Hard Stop Protocol**-the assessment must be immediately terminated if the participant exhibits disorientation or confusion, severely disorganized speech, acute psychosis, or paranoia, or if they are acutely under the influence of illicit drugs or severely intoxicated to the point of preventing understanding. The assessment will also cease if there is any verbal or non-verbal indication of distress.

**Hypertensive Crisi**s-a critically elevated blood pressure reading that is greater than or equal to 180mmHg systolic and greater than 120mmHg diastolic values.

**Inclusion Criteria-**to be eligible for participation, individuals had to be over the age of 18 and currently unsheltered, defined as residing in a place not meant for human habitation. Participants were required to demonstrate full cognitive capacity at the time of the encounter. This was verified by a standardized Alert and Oriented Assessment (A&O x4) of orientation to person, place, time, and event. Participants were required to provide explicit verbal consent after being informed of the assessment’s purpose and the screening’s voluntary nature.

**Numeric Pain Scale Rating (NPSR)** - a standardized 1-10 scale used in medical settings to quantify subjective reporting of pain. “1” serves as the benchmark for “no pain at all” and “10” serves as “the worst pain ever experienced.”

**Operational Needs Assessment**-a data collection activity performed solely to inform and improve the outreach efforts of a nonprofit organization (i.e., Valliant Foundation) or community-based outreach operations, distinct from formal research seeking a hypothesis-based conclusion.

**Red Flag Criteria-**thresholds for physiological metrics that trigger initiation of the Emergency Response Protocol, requiring immediate attention and offering of emergency services.

**Regular check-ins-**are scheduled meetings between the research team and the ethics committee to monitor study progress, ensure ongoing ethical compliance, address emerging issues, and maintain accountability throughout the research process.

**Unsheltered population**-any individual lacking a stable, regular, and adequate residence. Includes any individual living on streets, in vehicles, tents, temporary encampments, or shelter not belonging to them and not used for social purposes (i.e., government-funded hotel room)

## Appendix B: Field Narratives and Lessons Learned

The Mobile Outreach Initiative (MOI) was implemented to provide a respectful, consent-based health service and to connect unhoused individuals with essential resources, including free clinics, mental health and substance use services, city-provided shelters, and the Homeless Heartline operated by the Valliant Foundation. Beyond providing basic medical screening, the initiative’s effectiveness is grounded in personal connection, empathetic listening, and affirming participants’ dignity in each encounter.

During an outreach mission in the Mission District, a group of three volunteers encountered an individual who provided verbal consent for a free basic medical screening, including blood pressure, blood glucose, temperature, oxygen saturation, heart rate, and respiratory rate, as well as a public health questionnaire. This interaction was conducted through open dialogue and empathetic listening, ensuring the participant felt heard and respected. The team also provided a sheet detailing our foundation’s Homeless Heartline, free healthcare clinics for check-ups, and other community resources for the participant to use if they choose. This initial interaction demonstrates a high level of acceptability of the MOI’s services. A real-world demonstration of our person-centered approach was evident during a key moment of the encounter. The participant requested to use a team member’s phone to call his mother, expressing a basic human need for connection. Facilitating this request in a public health setting demonstrates both a simple gesture and a foundational principle: that humans have a right to a standard of living adequate for health, including social connection and care [7]. The conversation that followed was private, but afterward, the participant expressed feeling a sense of restored connection and dignity. The participant thanked the team, validating the power of our human-centered approach.

The effectiveness of this protocol was demonstrated when the team reencountered a participant in Union Square approximately 10 days after the initial assessment. The participant shared an update on their well-being, reporting that since the initial encounter, they have started reducing their smoking and are feeling better, with a sense of starting strong. Consistent with the program’s protocol, the team did not re-administer the formal assessment or record new physiological data; instead, the participant shared a verbal update on their well-being and smoking cessation efforts. Vital sign screenings may be offered, but data is never recorded a second time for the same participant. This ensured the participant received continuous, human-centered care without compromising the cross-sectional design.

During another outreach mission in the Tenderloin, a large group of unhoused individuals shared their concerns with multiple volunteers about high blood pressure readings and experiences with hypertension. The screener included a section to report difficulty finding outreach programs or primary care resources to help manage their condition. It was found that a majority of unhoused persons experienced difficulty gaining access to and maintaining continuity of care. Following MOI protocol, volunteers consulted with supervising medical professionals, listened attentively to the participants’ concerns, and reassured them that the free clinics listed on the foundation’s resource sheet provide access to healthcare services. In the lower part of the Tenderloin, volunteers offered extra food, water, and emergency blankets to unhoused individuals who declined the full screening but wanted a brief physical check-up and a conversation. Some participants told others about the volunteers’ presence to help the community, creating a safe environment for others to seek check-ins. Several provided verbal consent to complete both the physical screening and the questionnaire.

Volunteers conducted these interactions with care, aligned with program procedures, and made participants feel supported and cared for. As a result, many expressed gratitude for the team’s efforts, verbalizing that they felt treated with dignity and not overlooked. This feedback underscores the value of our outreach in supporting the unhoused community, which is often unheard.

While most interactions with participants do not result in an emergent need for medical attention, there have been incidents in which participants and volunteers encounter emotionally complex situations that underscore the importance of empathy, emotional validation, and respect for autonomy during crises and health emergencies. During an encounter with an unhoused individual, the conversation began when the participant noticed a cross necklace a volunteer was wearing. The participant, a native Spanish speaker, was able to communicate comfortably as volunteers proficient in Spanish engaged in conversation using the participant’s preferred language. Volunteers intentionally positioned themselves at eye level, adopted open, welcoming body language, and maintained a warm, attentive demeanor. This approach facilitated an atmosphere of respect and trust as the vital sign screening began. The participant shared that they had experienced homelessness for approximately 17 years, describing a profound faith that divine intervention had sustained and protected them throughout their time without shelter. The participant expressed a belief that God had safeguarded them throughout their experience of being unsheltered.

As dialogue between the volunteers and the participant continued, the conversation took an abrupt turn as the participant became tearful and expressed that they had been experiencing increasing suicidal ideation over the past year. During the interaction, volunteers adhered to the organization’s Emergency Response Protocol, systematically progressing through de-escalation measures while prioritizing the participant’s safety and autonomy. When the participant expressed a desire for professional assistance, volunteers facilitated activation of emergency medical services and coordinated ambulance transport in accordance with the participant’s request. Throughout this process, the volunteers maintained continuous engagement, provided reassurance, and monitored the participant’s emotional and physiological status to prevent escalation and ensure comfort. This experience highlights the importance of culturally and linguistically responsive care, which not only supports effective crisis management but also promotes trust, emotional security, and respect for participants’ self-determination.

Regarding other incidents in which the Emergency Response Protocol was utilized in MOI, EMS activation was warranted; however, participants’ autonomy was respected as they declined 911 transport. On two separate occasions during blood glucose checks using glucometers, participants had hypoglycemia. In both circumstances, the participants were A&Ox4 and informed that they had a blood glucose level that was potentially dangerous if a medical professional did not intervene.

On each occasion, both participants declined EMS activation for different reasons. One participant expressed concerns about the cost of an ambulance, fearing it would result in an unaffordable medical bill in addition to the cost of being seen at an emergency department, where complex levels of care are provided and can lead to expensive bills. The other participant also politely declined, expressing uncertainty and unease about going to the emergency room for undisclosed reasons. In both instances, the outreach volunteers respected participants’ wishes while ensuring they were informed of potential risks. Such interactions underscore the ethical balance between protocol obligations and honoring autonomy.

